# Investigating the Data Addition Dilemma in Longitudinal TBI MRI

**DOI:** 10.1101/2025.09.29.25336939

**Authors:** Antara Titikhsha, Mauluda Akhtar, AKM Moniruzzaman Mollah

## Abstract

Clinical machine learning (CML)for brain MRI often assumes that **more data guarantees better performance**, yet added samples can reduce accuracy when they arise from a different distribution, a phenomenon known as the **Data Addition Dilemma**. We present a systematic study of this issue in longitudinal TBI MRI, where acute baseline scans (S1) and follow-up scans (S2) differ substantially. Using a 14-subject, 28-scan cohort, we quantify the combined effects of intra-subject session shifts and inter-subject variability on severity classification. We evaluate four training schemes: (1) intra-session upper bound (S1→S1), (2) cross-session OOD testing (S1→S2), (3) pooled training (S1+S2→S1,S2), and (4) LOSO-IPA, which adds one unlabeled S2 scan per patient. With a lightweight logistic-regression model on PCA features, we show that naive pooling can degrade accuracy, pooled training trades baseline performance for modest robustness gains, and LOSOIPA recovers accuracy close to the intra-session limit. We recommend per-subject follow-up anchoring and diagonal CORAL alignment to mitigate session effects. These results clarify when additional data help or hinder CML workflows and provide a minimally invasive strategy for reliable longitudinal TBI severity assessment.

## 1 Introduction

In clinical machine learning for brain MRI, the assumption that larger datasets improve generalization breaks down when added samples differ from the original distribution. Even subtle demographic or scanner shifts can degrade performance, a phenomenon formalized as the *Data Addition Dilemma* [3]. This challenge is heightened in medical imaging, where multi-center variability and longitudinal changes between acute and follow-up TBI scans introduce substantial distributional drift [4, 5]. Using a 14-subject longitudinal cohort, we assess whether adding follow-up or cross-patient scans improves severity classification and disentangle intra- from inter-subject variability. Naive data addition offers no benefit and can impair performance, whereas targeted, distribution-aware strategies such as similarity-weighted augmentation or diagonal CORAL provide more reliable gains.

## 2 Contributions

This work makes the following contributions to clinical machine learning for traumatic brain injury (TBI) MRI:

1. **Problem framing: the Data Addition Dilemma**. We examine whether adding follow-up scans or cross-patient data improves or impairs generalization in TBI severity prediction, providing a clinically grounded instance of the broader *Data Addition Dilemma*.
2. **Longitudinal cohort analysis**. Using a compact 3D CNN trained on acute (S1) and follow-up (S2) MRI volumes, we disentangle intra-subject shifts from inter-subject variability and show that naive data addition does not reliably enhance performance.
3. **Evaluation of augmentation strategies**. We assess baseline training, LOSO-IPA, pooled training, cross-patient augmentation, and diagonal CORAL, identifying when augmentation helps, when it fails, and when alignment is essential.
4. **Interpretability via dimensionality reduction**. PCA and t-SNE projections reveal session- and severity-related structure in the latent space, highlighting how distribution shifts appear across patients and timepoints.
5. **Implications for deployment**. Our results caution against assuming that *more data = better performance* in heterogeneous MRI settings and instead support minimal per-subject adaptation with domain alignment as a more reliable path toward robust TBI severity modeling.

## 3 Prior Works

### 3.1 The Data Addition Dilemma in Machine Learning

Classical learning theory suggests that larger datasets should improve generalization [1, 2], yet Shen et al. showed that adding OOD samples can *decrease* performance under distributional shift [3]. This Data Addition Dilemma is especially concerning for clinical ML, where heterogeneity is unavoidable and naive pooling can disadvantage underrepresented groups [7].

### 3.2 Distribution Shifts in Medical Imaging

Medical imaging routinely encounters substantial distributional variability [2, 4]. Multi-center datasets differ in scanners, field strengths, and acquisition protocols [8], and even within a single subject, temporal changes between acute and follow-up TBI scans alter intensities and motion patterns [5, 9]. These shifts undermine the assumption of matched training and testing distributions.

### 3.3 Longitudinal TBI Studies

Longitudinal TBI MRI reveals evolving neuroanatomical and diffusion patterns [9, 10], making temporal labels inconsistent as severity can improve or worsen over time. Treating follow-up scans as interchangeable training samples risks encoding these confounding dynamics, motivating approaches that explicitly model temporal change rather than pooling sessions indiscriminately.

### 3.4 Domain Adaptation and CORAL

Domain adaptation provides a principled way to address distribution mismatch in medical imaging [11]. CORAL offers a simple and efficient solution by aligning second-order statistics between source and target domains [6], and its variants have proven effective across multiple MRI modalities [12].

### 3.5 Leave-One-Subject-Out Validation

LOSO cross-validation remains a gold standard for neuroimaging studies [13]. By withholding all scans from each subject in turn, LOSO avoids data leakage and yields a rigorous measure of generalization, particularly important for small clinical cohorts such as TBI [14].

### 3.6 Out-of-Distribution Detection

OOD detection aims to identify samples outside the training distribution [15], but many uncertainty-and reconstruction-based methods perform poorly on MRI [16]. This has motivated new clinical OOD frameworks designed specifically for safety-critical medical-imaging settings [17].

## 4 Methods

### 4.1 Dataset

We analyzed a pilot cohort of 14 patients with traumatic brain injury from OpenNeuro (ds00022) [18], spanning mild, moderate, and severe injury cases. Each subject contributed two MRI sessions: an acute baseline scan (S1) and a follow-up scan (S2), yielding 28 volumes. All scans were preprocessed into standardized 64 × 64 × 64 grids, normalized, and prepared for downstream modeling.

### 4.2 Experiment 1: Naive Augmentation with a 3D CNN

To assess whether follow-up scans improve or degrade severity classification, we trained a compact 3D CNN (shown in Figure 1)composed of sequential 3 ×3 × 3 convolutional blocks with batch normalization and max pooling, followed by global average pooling and a fully connected softmax head.

**Fig. 1.**
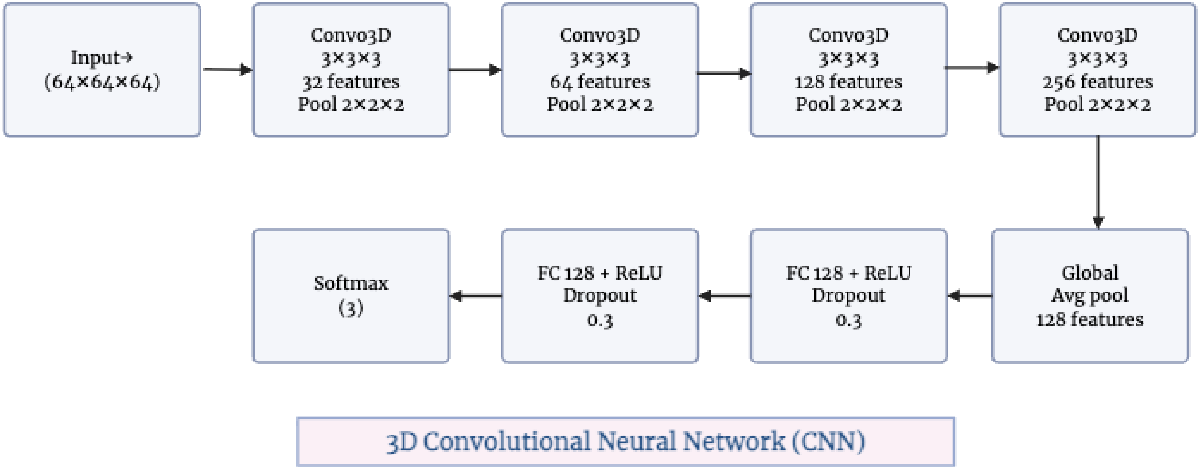
3D CNN architecture used in Experiment 1.

Shown in Table 1, we evaluated four augmentation strategies under identical training settings (Adam, early stopping, cross-entropy). The baseline trained only on S1 and tested on S2 to measure cross-session performance. Geometric augmentation applied flips, small rotations, and elastic deformations to S1 scans. Follow-up augmentation added each subject’s S2 scan as an independent sample. Cross-patient augmentation incorporated S2 scans from other subjects. This experiment examines how naive inclusion of additional scans influences model robustness in a longitudinal TBI setting.

**Table 1.** Performance comparison of augmentation strategies on the held-out S2 test set.

| Strategy | Accuracy | F1-macro |
| --- | --- | --- |
| Baseline | 64.3% | 0.261 |
| Geometric | 14.3% | 0.083 |
| Follow-up | 64.3% | 0.253 |
| Cross-patient | 57.1% | 0.367 |

### 4.3 Experiment 2: Probing the Data Addition Dilemma with PCA and Logistic Regression

To address the limitations of naive augmentation in **Experiment 1**, we adopted a transparent, low-capacity pipeline grounded in the *Data Addition Dilemma*. Each MRI volume was resampled to 64 × 64 × 64, vectorized, and reduced to 50 principal components to retain dominant variance while limiting overfitting. A multinomial logistic-regression model, chosen for its interpretability and suitability for small datasets, was evaluated under four data-addition regimes: **(i)** intra-session upper bound (S1→ S1), **(ii)** cross-session OOD test (S1→ S2), **(iii)** pooled training (S1+S2 → S1,S2), and **(iv)** LOSO-IPA, which adds one unlabeled S2 scan per patient within a leave-one-subject-out framework. Accuracy and macro-F1 were used to assess class-balanced performance across mild, moderate, and severe TBI. This experiment clarifies when additional scans improve generalization and when they amplify distributional mismatch.

### 4.4 Experiment 3: Domain Adaptation and Visualization

To address the limitations of naive augmentation (Experiment 1) and linear modeling (Experiment 2), we applied domain adaptation methods to reduce distributional mismatch between acute (S1) and follow-up (S2) embeddings within the logistic-regression pipeline.

#### Domain Adaptation Methods

i. **Diagonal CORAL**. We aligned second-order statistics by re-scaling each PCA dimension of S1 to match the variance of S2, providing an efficient way to mitigate session-induced shifts.
ii. **Similarity-Weighted S2 Augmentation**. We computed cosine similarity between each S2 embedding and the S1 centroid, assigning higher weight to more similar follow-up samples and down-weighting outliers to retain useful information without amplifying session noise.

#### Visualization of Adaptation Effects

PCA and t-SNE projections were used to illustrate how adaptation reshapes the feature space, including **(i)** reduced separation between S1 and S2 distributions and **(ii)** clearer severity-related structure after alignment.

#### Evaluation Metrics

Accuracy and macro-F1 quantified overall and class-balanced performance. Together, these analyses show that principled domain adaptation and selective augmentation substantially reduce cross-session mismatch in longitudinal TBI MRI.

## 5 Results and Discussion

### 5.1 Experiment 1: Naive Augmentation with a 3D CNN

The baseline S1→S2 model achieved 64.3% accuracy and a macro-F1 of 0.261, reflecting the substantial shift between acute and follow-up scans. Geometric augmentation sharply reduced performance, indicating that standard perturbations obscure subtle injury-related structure in TBI MRI. Adding each subject’s S2 scan offered no improvement, showing that intra-patient follow-ups—affected by healing and scanner drift—do not enhance cross-session generalization. Cross-patient augmentation increased F1 but lowered accuracy, suggesting gains for minority classes at the cost of added heterogeneity. Overall, naive augmentation fails to resolve the session mismatch and can worsen classification.

Figure 2 shows that baseline performance is concentrated in the Severe class, geometric augmentation offers minimal improvement, follow-up augmentation primarily boosts Severe predictions, and cross-patient augmentation improves Mild and Moderate recognition at the expense of Severe.

**Fig. 2.**
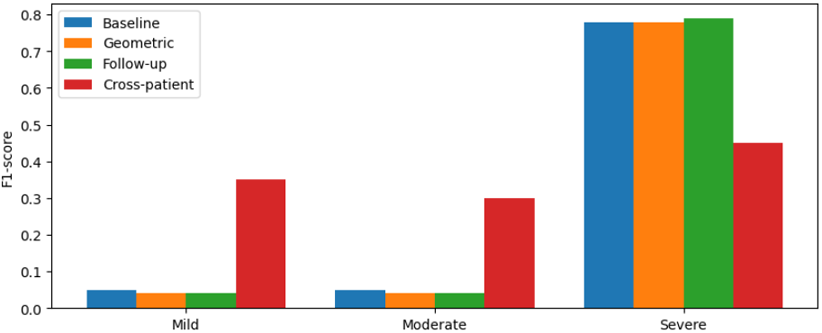
Class-level F1 scores across augmentation strategies.

Figure 3 shows that additional scans modestly increase dataset size, yet these increases do not translate into performance gains, illustrating the Data Addition Dilemma.

**Fig. 3.**
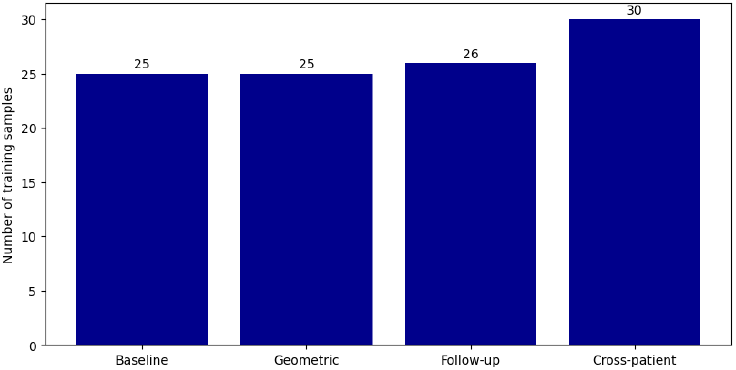
Training sample counts for each augmentation strategy.

Figure 4 shows that, despite incorporating more samples in follow-up and cross-patient regimes, performance does not improve, suggesting that heterogeneous additions introduce noise rather than a useful signal.

**Fig. 4.**
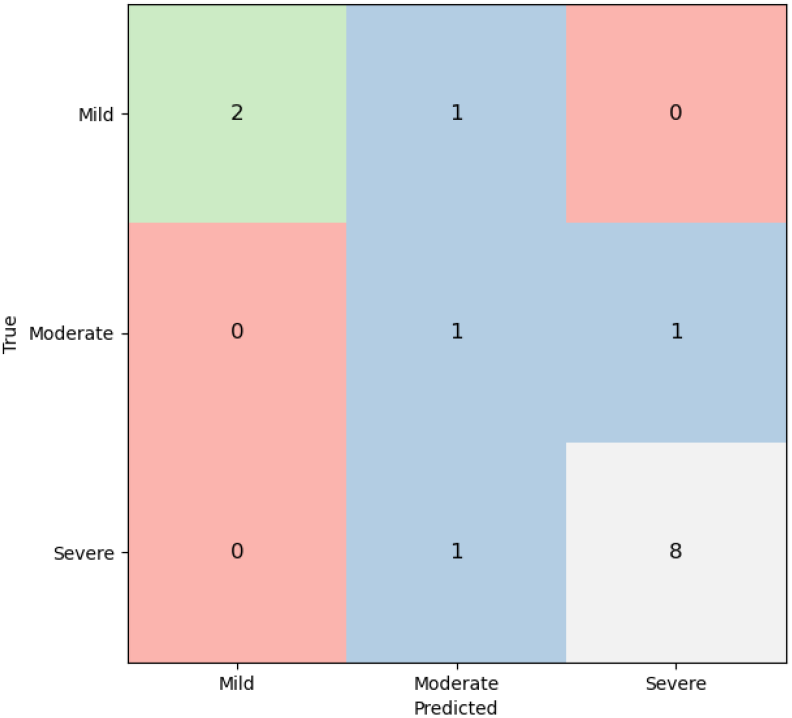
Number of training samples by augmentation strategy.

Figure 5 shows that Severe cases dominate predictions, while all Mild and Moderate cases are misclassified, highlighting strong session-induced drift and limited separability.

**Fig. 5.**
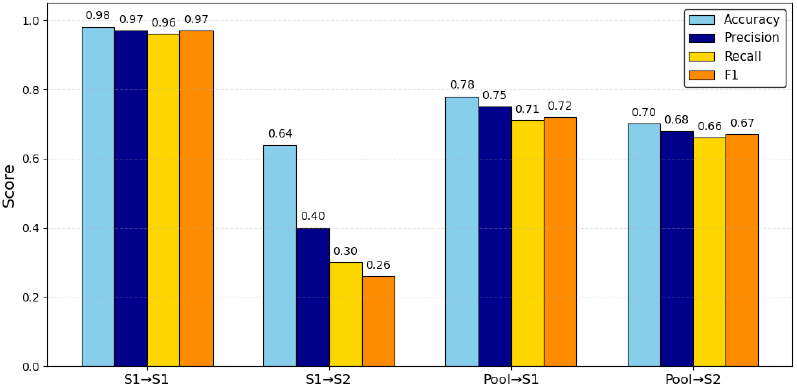
Baseline confusion matrix for S1-trained, S2-tested model.

### 5.2 Experiment 2: Data Addition Dilemma with PCA and Logistic Regression

Using 50-component PCA embeddings, logistic regression reached its highest performance under S1 → S1 evaluation, confirming that severity structure is recoverable when distributions match. Cross-session S1 → S2 performance dropped sharply due to session effects such as tissue recovery and scanner variation. Pooled training did not improve robustness and diluted useful discriminative cues. LOSO-IPA, which adds one unlabeled S2 scan per subject, yielded modest F1 gains, showing that small amounts of subject-specific adaptation help more than naive pooling. These results emphasize that alignment and relevance of added data are more important than quantity.

Figure 6 reveals substantial confusion across severity levels in cross-session evaluation.

**Fig. 6.**
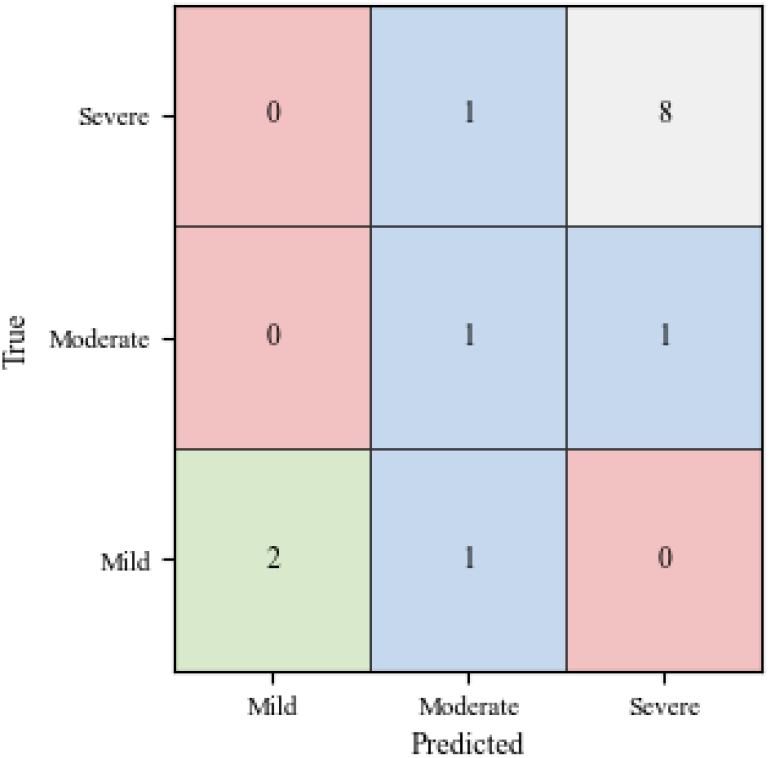
Performance across training–testing regimes.

### 5.3 Experiment 3: Domain Adaptation and Visualization

Diagonal CORAL reduced S1–S2 variance mismatch and improved accuracy and macro-F1, particularly for Moderate and Severe classes. Similarity-weighted augmentation further balanced class performance by emphasizing S2 scans most consistent with S1. PCA and t-SNE projections showed that adaptation reduced session separation and clarified severity structure, whereas unadapted embeddings clustered primarily by session. Together, these results show that targeted domain alignment, not naive data addition, is necessary to recover clinically meaningful structure in longitudinal TBI MRI.

Figure 7 shows uneven class performance: Severe cases are easiest to identify, Mild cases partially recoverable, and Moderate cases most ambiguous due to embedding overlap.

**Fig. 7.**
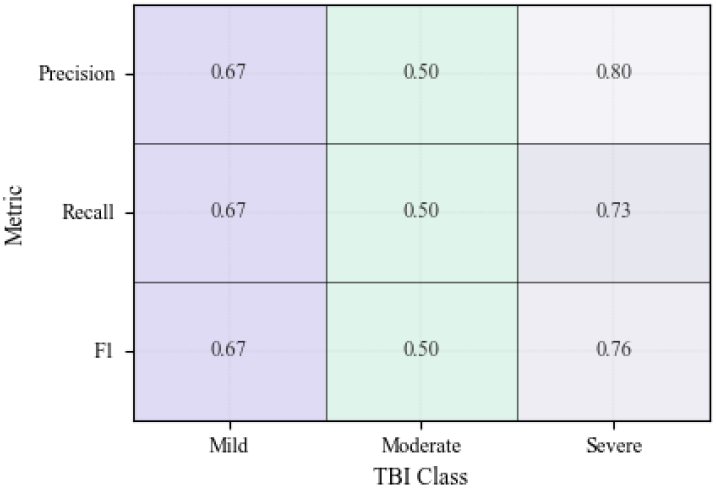
Per-class classification performance.

Figure 8 shaows macro-precision, recall, and F1 remain moderate but balanced, indicating that errors stem from broad class overlap rather than systematic bias.

**Fig. 8.**
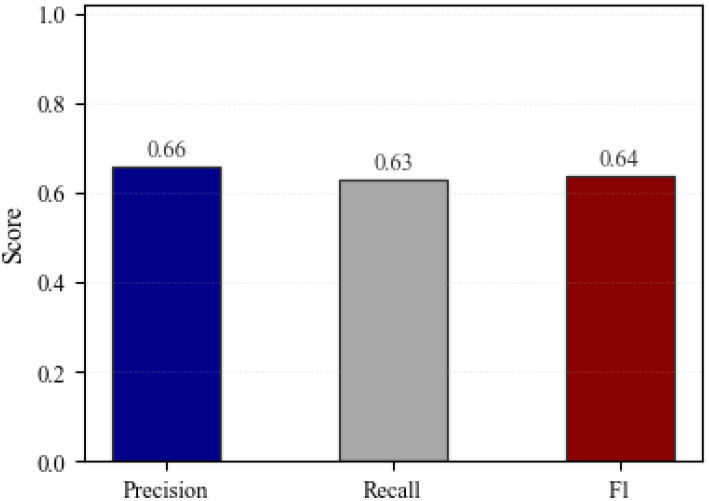
Macro-averaged metrics after adaptation.

Figure 9 shows substantial overlap in unadapted embeddings and improved but still partial class separation after domain adaptation, underscoring the subtlety of the severity signal relative to session and subject variability.

**Fig. 9.**
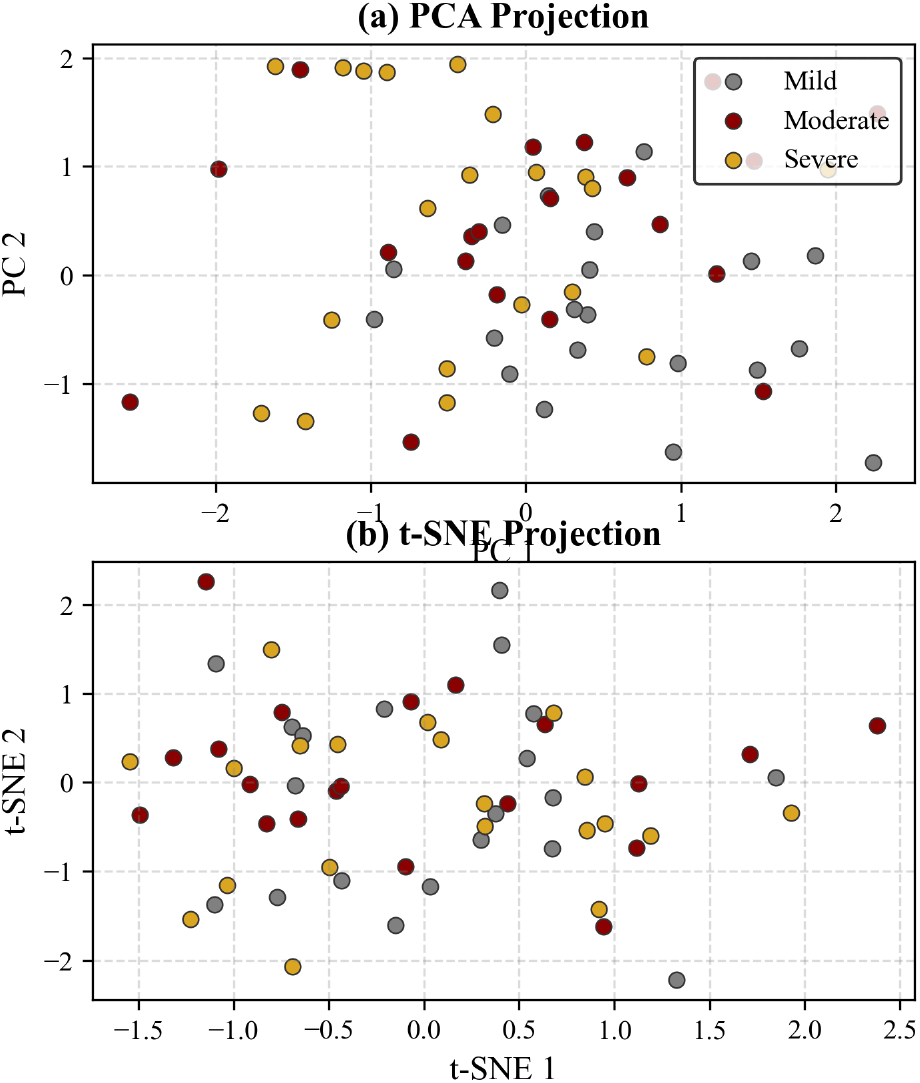
PCA and t-SNE projections before and after adaptation.

## 6 Limitations and Future Work

This pilot study is limited by its small cohort, evolving severity labels, and the use of simple classifiers, as well as the absence of demographic and clinical metadata for subgroup or fairness analysis. Future work will explore stronger domain-adaptation methods and longitudinal models, along with distribution-aware sample selection and anatomically informed augmentation. Integrating clinical and demographic variables will further enable subgroup evaluation and support equitable model performance.

## 7 Conclusion

This pilot study examined the Data Addition Dilemma in longitudinal TBI MRI and showed that naive pooling does not resolve session mismatch and can hinder classification. Across both CNN and PCA–logistic pipelines, minimal per-subject adaptation and lightweight alignment methods (diagonal CORAL, similarity-weighted augmentation) offered the most reliable improvements. Future work will explore richer adaptation frameworks and explicit longitudinal models, alongside principled sample selection and anatomically informed augmentation. Incorporating demographic and clinical metadata will further enable subgroup and fairness analyses. Together, these directions outline a concise path toward robust and equitable longitudinal TBI severity assessment.

## Data Availability

The dataset has been downloaded from here: https://openneuro.org/datasets/ds000220/versions/00002

## Acknowledgements

We thank Sidhika Balachandar for her invaluable feedback and discussions, and Divyanshu Tak for his essential contributions to MRI preprocessing. We are also grateful to members of the Wood Neuro Research group for their mentorship and insights. Finally, we acknowledge Roy et al. (2017) for publicly releasing the ds00022 dataset [18].

